# Exploratory analyses of Immunologic Features in a Randomized, Placebo-Controlled Trial of Nirmatrelvir/Ritonavir for Long COVID

**DOI:** 10.64898/2026.02.24.26347001

**Authors:** Bornali Bhattacharjee, Mitsuaki Sawano, William B. Hooper, Kexin Wang, Alexandra Tabachnikova, Valter Silva Monteiro, Peiwen Lu, Pavlina Baevova, Gisele C. Rodrigues, Victoria L. Fisher, César Caraballo, Rohan Khera, Shu-Xia Li, Jeph Herrin, Dany Christian, Andreas Coppi, Frederick Warner, Julie Holub, Yashira Henriquez, Maria A. Johnson, Theresa B. Goddard, Erica Rocco, Amy C. Hummel, Mohammad AL Mouslmani, Kevin D. Carr, Lawrence Charnas, Magdia De Jesus, Dale Nepert, Paula Abreu, Frank W. Ziegler, John A. Spertus, Leying Guan, Harlan M Krumholz, Akiko Iwasaki

## Abstract

This exploratory analysis of PAX LC, a Phase 2, 1:1 randomized, double-blind, superiority, placebo-controlled trial examined whether treatment with nirmatrelvir/ritonavir (NMV/r) versus placebo/ritonavir (PBO/r) in individuals with Long COVID could reveal immune features associated with symptom improvement. Eighty-two participants (n=45 PBO/r; n=37 NMV/r) provided blood samples at baseline (Day 0) and post-treatment (Day 28). Baseline demographic and immunological phenotypes were similar in the two groups. No significant differences were observed in major immune cell populations or organ function markers between NMV/r vs. PBO/r groups, or before vs. after the treatment. Modest hematologic changes were noted in the NMV/r arm. SARS-CoV-2-specific IgG levels remained constant, with changes in total immunoglobulin subtypes and isotypes in both arms. Both arms showed similar shifts in cytokine levels. Notably, the levels of S1 and Spike proteins in circulation remained unchanged post-treatment. Regardless of the treatment arm, participants with self-reported symptom improvement showed reductions in the level of the inflammatory chemokine RANTES. Taken together, the findings of this study demonstrate limited virological and immunological changes in response to nirmatrelvir, contributing insights into the reason for the lack of benefit of the 15-day NMV/r treatment in Long COVID.

## Introduction

Viral persistence is proposed as one of the key drivers of Long COVID^1^. However, the spectrum of clinical manifestations and multi-organ involvement suggests that multiple overlapping mechanisms may be at play, leading to different disease patterns^2,3,4^. This heterogeneity adds significant complexity to both diagnosis and treatment. Additionally, it has been challenging to detect persistent viral presence from accessible biospecimens such as blood or other easily collectible body fluids. Part of the challenge lies in the fact that the virus may reside in tissues without shedding sufficient material into the bloodstream to be detectable. Despite these challenges, several clinical trials are actively investigating whether clearing residual virus can alleviate Long COVID symptoms^5^. These include the antiviral ensitrelvir (NCT06161688; PREVAIL-LC), the gut-targeting agent larazotide (NCT05747534), and the monoclonal antibody AER002 (NCT05877508), among others. The antiviral agent, Paxlovid (nirmatrelvir (PF-07321332)/ritonavir; NMV/r)^6^ has been evaluated across four Long COVID trials (NCT05576662; STOP-PASC, NCT05823896; PROLIFIC, NCT05595369; RECOVER-VITAL), including one which was conducted by our team (NCT05668091; PAX LC). Together, these trials represent a critical step toward determining whether antiviral treatment can improve the health of people suffering from Long COVID.

The first Paxlovid trial to be completed was the STOP-PASC trial conducted at Stanford University^7^. It was a randomized, placebo-controlled study evaluating the efficacy of a 15-day course of NMV/r in 155 participants with Long COVID symptoms lasting over three months. The primary endpoint comprised of changes in combined severity score of six common symptoms, including fatigue, brain fog, and shortness of breath, but did not show any statistically significant difference between the nirmatrelvir and placebo groups at 10 weeks. Along similar lines, the PAX LC trial, conducted by our team at Yale^8,9^, evaluated the same 15-day NMV/r regimen in 100 participants. Unlike the earlier single-site study, this trial was fully decentralized, enrolling participants from 28 states across the U.S. The primary outcome was a change in the PROMIS-29 Physical Health Summary Score at day 28, which did not differ significantly between the NMV/r and placebo/ritonavir (PBO/r) arms of treatment.

In alignment with one of the exploratory objectives of the PAX LC trial, we examined immunological and virological features in the blood of participants before and after the treatment course. The three objectives of this sub-study were: 1. to evaluate if there were baseline immunological differences between the cohorts in the two treatment arms; 2. to investigate whether treatment resulted in changes in the immune and circulating viral spike protein in participants; and 3. to determine whether symptom trajectories could be explained by changes in the levels of immunological factors, irrespective of treatment allocation. We used multiple orthogonal approaches to measure cytokines, antibodies, and peripheral blood mononuclear cell phenotypes.

## Results

### Participant characteristics

Individuals with Long COVID from 28 of the 48 contiguous US states were enrolled between March and August 2024. A total of 45 from the PBO/r arm and 37 from the NMV/r arm were included in this exploratory immunophenotyping study (Extended data fig 1, Fig 1A). The groups were similar in age (median 42 vs. 43 years; Mann-Whitney U test p_adj_= 0.7366), sex distribution (64.4% vs. 73% female; Chi-square test p_adj_= 0.5452), vaccination history (median four doses at baseline) and duration of Long COVID symptoms (72–1392 days; Mann-Whitney U test p_adj_ = 0.2072). As Long COVID outcomes may also be modulated by factors such as vaccination status and infection with viral variants among others^10^, participants in each arm were further stratified by viral variant waves on the basis of self-reported index infection timing. The participants’ self-reported COVID-19 index infection dates were matched by analyzing the variant sequencing data from the GISAID database^11^, and the WHO variant wave classification to assign variants. A higher proportion of participants in the PBO/r arm were infected during early variant waves [PBO/r: n= 18 (40%); NMV/r: n= 5 (13.5%); Chi-Square test p_adj_= 0.0632; Fig 1C] and were infected before completing the primary series of vaccination [PBO/r: n= 20 (44.4%); NMV/r: n=9 (24.3%); Chi-Square test p_adj_= 0.2072; Fig 1D] but these differences did not reach significance.

**Fig 1:**
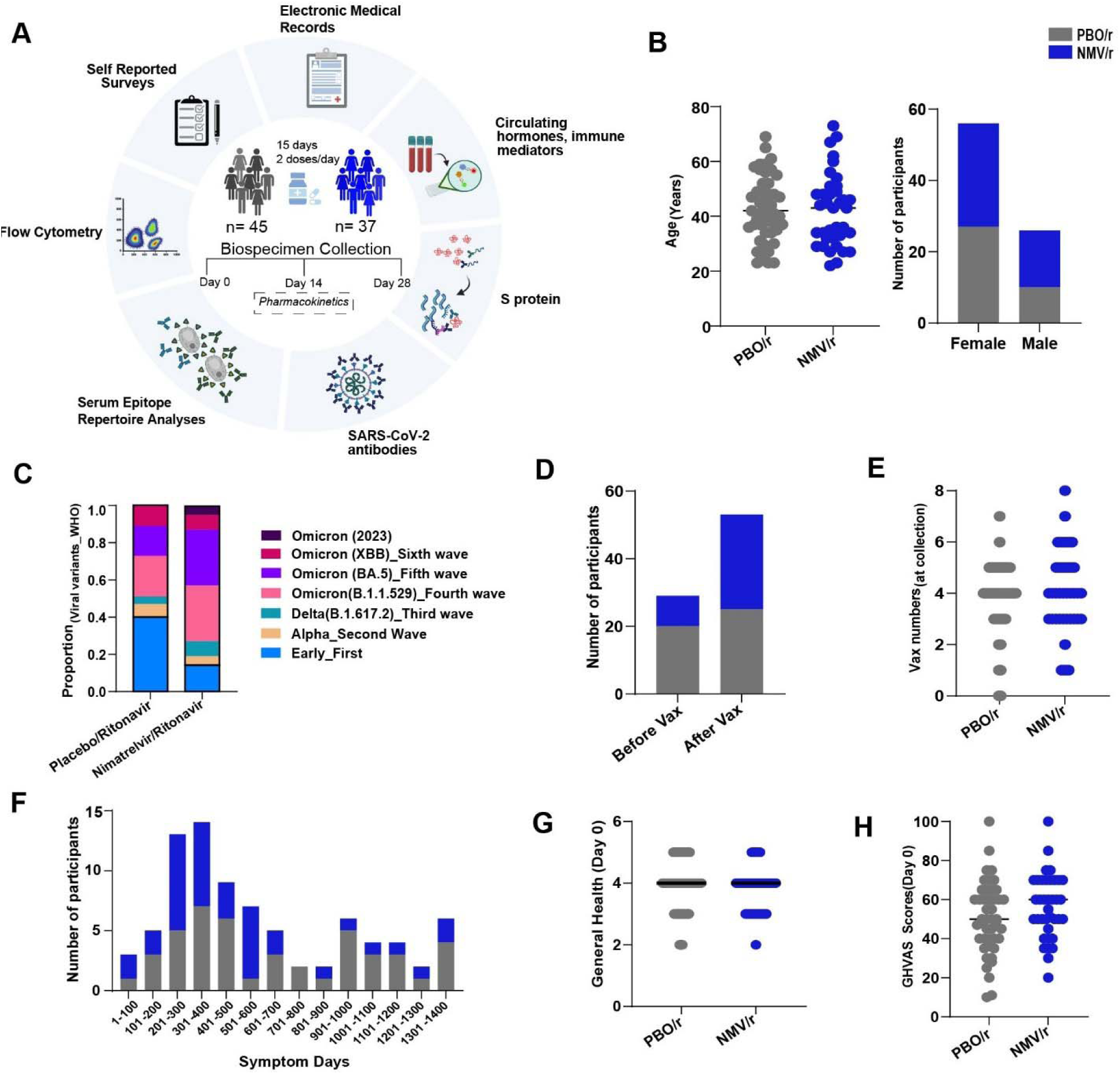
Study Design and Cohort Characteristics. **A.** Summary of treatment arms (PBO/r: n= 45; NMV/r: n= 37), study design and assays included in the study **B.** Age and biological sex breakdown by treatment group **C.** Stacked bar plots of index viral variants based on self-reported infection dates and variant waves **D.** Stacked bar plots indicating the number of participants in each arm based on index infection timing relative to primary series of COVID-19 vaccines [Before vaccination (Before vax), after vaccination (After vax)]. There were two non-vaccinated participants who were also included in the before vax arm. **E.** Scatter plots of vaccine numbers in the two treatment arms. **F.** Stacked bar plots of self-reported symptom days **G.** Boxplots showing self-reported general health status and General Health Visual Analogue Scale (GHVAS) scores self-reported at baseline. The p-values were calculated using the Mann Whitney U-tests for continuous variables and Chi-squared tests for categorical variables with Benjamini Hochberg correction. No statistically significant differences were observed.

Both groups had a median of four vaccine doses at baseline, with no difference in total doses (Mann-Whitney U padj = 0.8094; Fig 1E). Two participants in the placebo arm reported never receiving any COVID-19 vaccination. The duration of Long COVID symptoms (72 to 1392 days), measured as days from self-reported symptom onset to biospecimen collection, were similar between treatment arms (Mann-Whitney U test p_adj_ = 0.2072, Fig 1F). Baseline general health status (p_adj_ = 0.4851) and median General Health Visual Analogue Scale (GHVAS) scores (p_adj_ = 0.2348; Fig 1G) were also similar between groups.

### 15-day NMV/r treatment was associated with modest hematologic changes without affecting general blood cell counts in research assays

As part of the immunophenotyping analyses, complete blood counts were measured at baseline and day 28 using assays not intended for clinical use. No differences were observed between arms at baseline or day 28, and no within-group changes were detected in the placebo arm. In the NMV/r arm, modest but statistically significant shifts were detected, including reduced median red blood cell numbers, platelet counts, and hemoglobin levels, alongside increases in red cell distribution width and mean corpuscular volume (p_adj_ values 0.0027–0.0350; extended data table 2). Importantly, all values remained within the expected physiological range.

### Paxlovid treatment did not alter the SARS-CoV-2 Spike protein levels in circulation

The use of nirmatrelvir, an antiviral agent that targets the SARS-CoV-2 main protease, in Long COVID is grounded on the hypothesis that SARS-CoV-2 continues to replicate *in vivo*, leading to innate and adaptive immune responses as well as pathologies associated with the viral proteins^12^. Hence to assess the proportion of participants with viral persistence and assess the reduction of the same upon treatment, the presence of viral spike protein in circulation at baseline and day 28 were measured. Successive Proximity Extension Amplification Reaction (SPEAR) immunoassays were implemented for detection of S1 and full-length Spike (S) protein^13^. The lowest functional limit of detection (FLLoD) for the S1 assay was 7.04 femtomolar (fM) and that of the full-length S assay was 2.3 fM. The number of participants with detectable S1 was 25 (55.6%) and 16 (43.2%), respectively, in the PBO/r and the NMV/r arms at baseline. This difference was not statistically significant (Chi-Square test p= 0.2672; Fig. 2A).

**Fig 2:**
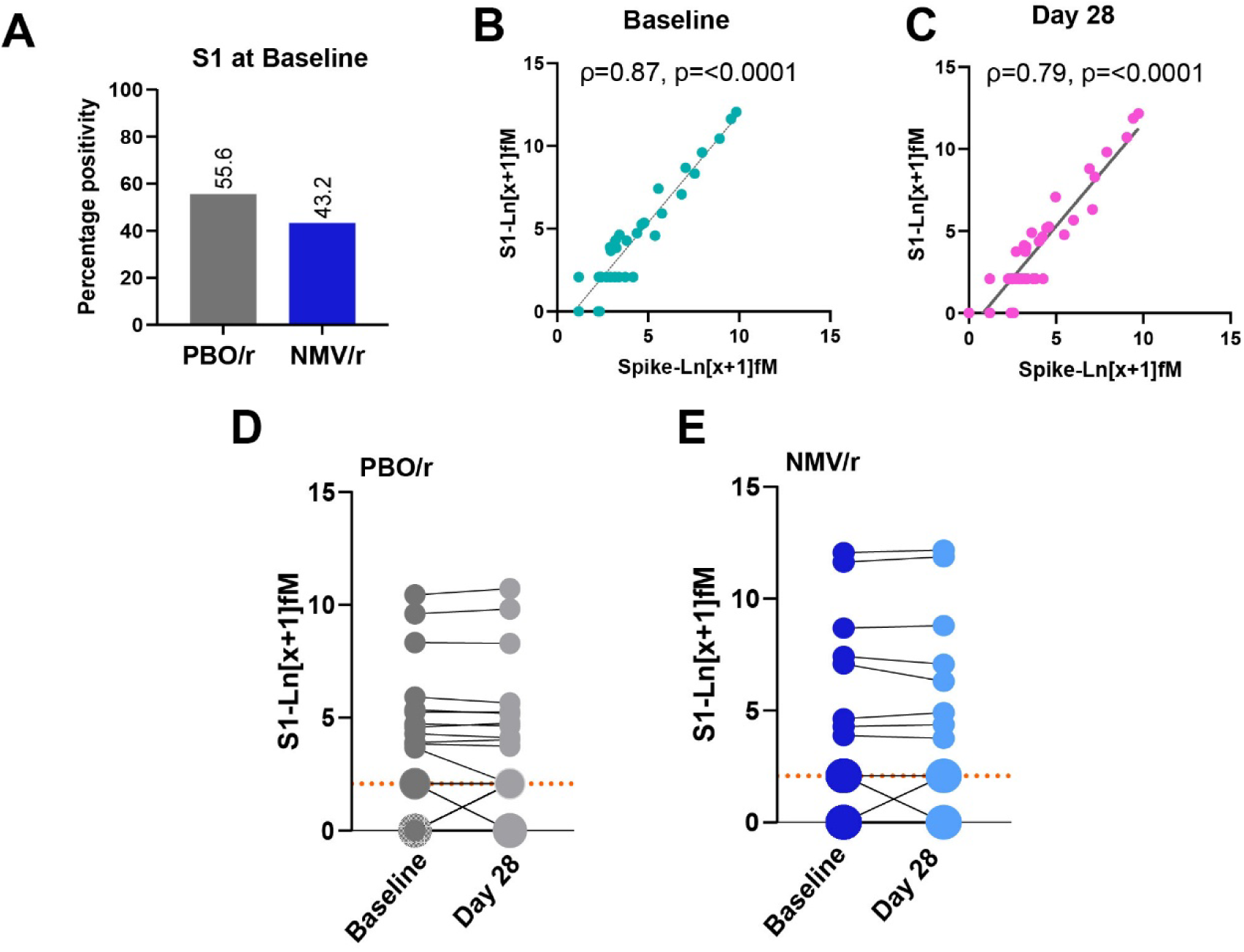
No differences were observed in circulating SARS-CoV-2 Spike protein levels after treatment. **A.** Bar plots indicate the percentage of participants with detectable circulating plasma S1 at baseline, with percentage values mentioned above each bar. No significant differences in positivity were observed between the two treatment arms based on Chi Square test **B-C.** Linear regressions of S protein levels as detected using the S1 and the full-length SPEAR assays at baseline and at day 28 respectively, irrespective of treatment arms. Spearman’s correlation was calculated with corresponding p-values. Dotted lines depict linear regressions. **D.** Paired line plots of circulating plasma S1 levels at baseline and day 28 in the PBO/r group **E.** Paired line plots of circulating plasma S1 levels at baseline and day 28 in the NMV/r group arm. The orange dotted lines indicate the LLoD. To evaluate differences between timepoints within each treatment arm, Wilcoxon matched-pairs signed-rank test was used. No statistically significant differences were observed. Larger circle size indicates values below the LLoD or LLoQ in multiple participants

To investigate whether the presence of S1 reflects the presence of full-length S protein and to validate the S1 findings, we next ran the full-length (ectodomain) S protein SPEAR assays in a separate batch. Significant correlations between S1 and S levels were observed from all samples at both baseline (Spearman Correlation ρ= 0.87, p= <0.0001; Fig. 2B) and on day 28 (Spearman Correlation ρ= 0.79, p= <0.0001, Fig. 2C), irrespective of drug usage, demonstrating high concordance between these assays. Furthermore, the S1 measurements mostly reflect the presence of the full-length ectodomain of the S protein, including the S2 domain.

To understand if Paxlovid caused changes in the levels of circulating S1, a surrogate marker for SARS-CoV-2 persistence^14^, baseline and day 28 S1 levels were compared using Wilcoxon matched-pairs signed-rank test. Similar analyses were also run in the PBO/r arm to exclude randomness. No significant differences were observed in either the PBO/r arm (p= 0.1468, Fig. 2D, extended data table 1) or the NMV/r arm (p= 0.9580, Fig. 2E, extended data table 1). These data indicate that the 15-day course of Paxlovid did not impact the levels of SARS-CoV-2 spike protein, as measured by the SPEAR assays.

### Paxlovid treatment does not alter SARS-CoV-2 specific antibody responses

A consistent feature across Long COVID studies has been the presence of elevated levels of circulating anti-Spike immunoglobulin G (IgG)^15–18^ despite reduced neutralizing abilities^19^. More recently, anti-RBD-specific IgG responses have been proposed as a sensitive marker of Long COVID, potentially linked to ongoing viral persistence^18^. To characterize humoral immune changes, plasma levels of total IgM, IgG subclasses (1, 2, 3, 4), and IgA were first assessed using a multiplex Luminex assay. Paired comparisons showed significant, but small increases in all immunoglobulin isotypes and IgG subclasses following treatment in the placebo arm (Fig. 3A, extended data table 1). In the NMV/r arm, similar increases were observed across all isotypes and IgG subclasses, except for IgG4 (Fig. 3B, extended data table 1).

**Fig 3:**
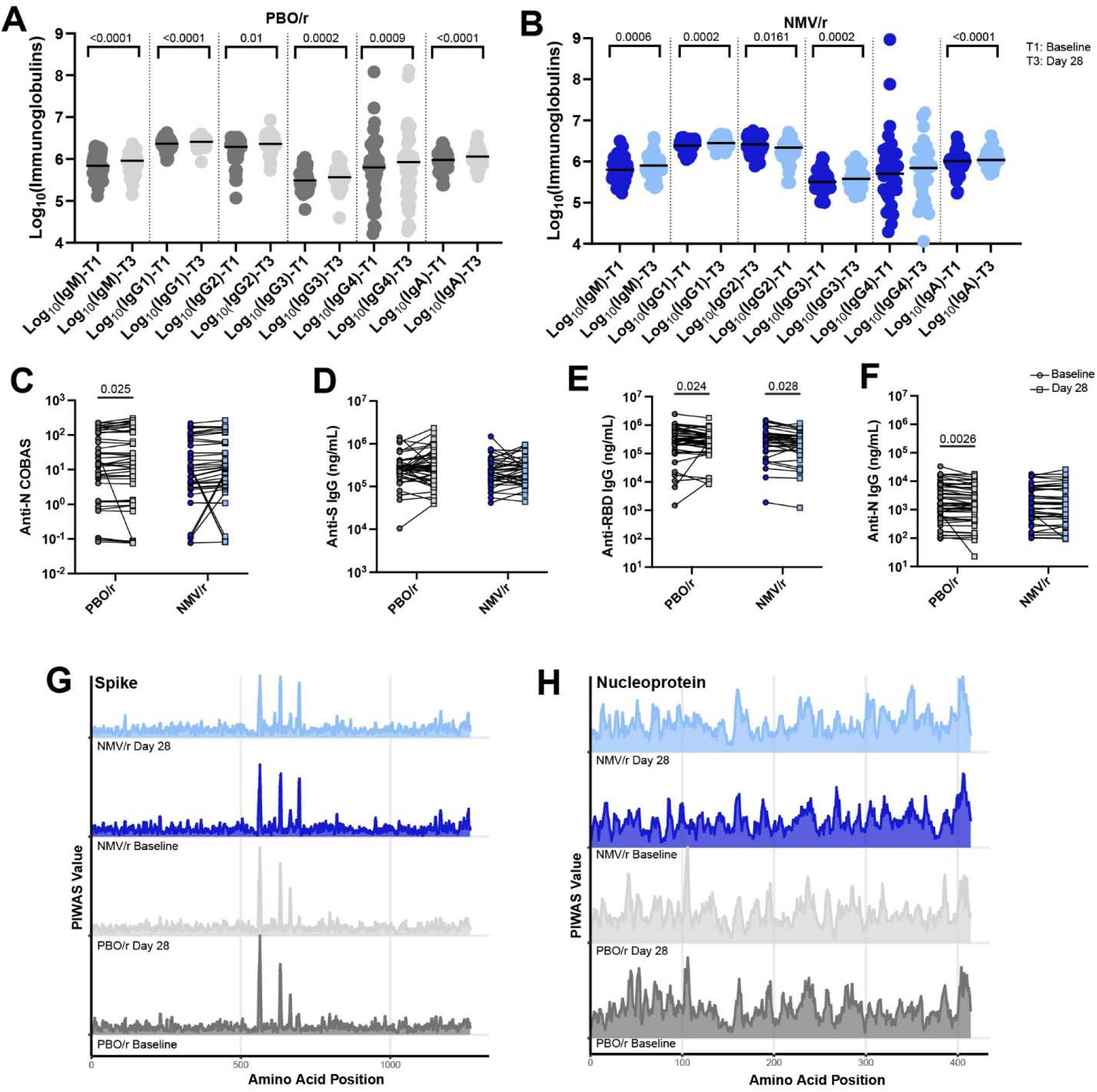
Antibody levels increase from baseline at Day 28 but are not indicative of an increase in anti-SARS-CoV-2 humoral responses. A-B. Log-transformed circulating total antibody subtypes and IgG isotypes, as measured by Luminex assays at baseline and day 28 in the two treatment arms. SARS- CoV-2 antibody responses were assessed using **C.** Nucleocapsid by COBAS assay, ELISA for **D**. Spike, **E**. Receptor Binding Domain (RBD), **F**. Nucleocapsid. Significance for difference in group median values was assessed using Wilcoxon matched-pairs signed-rank test with Benjamini–Hochberg false-discovery rate (FDR) correction for multiple comparisons. PIWAS line profiles of IgG binding within participants along the SARS-CoV-2 **G.** Spike and **H.** Nucleocapsid amino acid sequences. 95th percentile values of binding are arranged by group. P-values are provided for statistically significant differences.

To evaluate high-affinity anti-N antibody levels including IgG, sensitive EUA-cleared Elecsys anti-SARS-CoV-2 immunoassays were performed. A total of 12 participants had antibody indices below 1 at both time points (PBO/r= 8; NMV/r= 4). A significant increase (Wilcoxon Rank Sum test; p_adj PBO/r_ = 0.0462, p_adj NMV/r_ = 0.0254; Fig. 3C) in the placebo arm, with a median increase in antibody index of 0.18 at day 28, was observed. SARS-CoV-2-specific IgG responses were also evaluated using in-house enzyme-linked immunosorbent assays (ELISAs) targeting full-length Spike (S), the receptor-binding domain (RBD), and the Nucleocapsid (N) protein. No significant differences in anti-Spike IgG levels were observed in either arm (Wilcoxon Rank Sum test; p_adj PBO/r_ = 0.4409, p_adj NMV/r_ = 0.5605; Fig. 3D). However, a significant decline in RBD-specific IgG was observed on day 28 compared to baseline, irrespective of drug arms (Wilcoxon Rank Sum test; p_adj PBO/r_ = 0.024, p_adj NMV/r_ = 0.0275; Fig. 3E), with a small decrease of <0.5 log₁₀ in median titers at day 28. Similarly, significant differences were detected in anti-N IgG levels in the placebo arm, with the decline in median titers being <0.1 log₁₀ at day 28 (Wilcoxon Rank Sum test; p_adj PBO/r_ = 0.0026, p_adj NMV/r_ = 0.8137; Fig. 3F). These inconsistent findings and the very small changes observed make the interpretation of the anti-N IgG values difficult.

To further evaluate if there were differences in antibody epitopes against the SARS-CoV-2 S and N proteins, profiling was carried out for anti-SARS-CoV-2 IgG responses using linear peptides in vaccinated individuals using the (Protein-Based Immunome Wide Association Studies) PIWAS method^20^. Higher PIWAS values were observed for the dominant epitopes without differences between and within cohorts for both S (Fig. 3G) and N protein (Fig. 3H). Moreover, no ritonavir or nirmatrelvir-associated shifts were observed at day 28 (Fig. 3G, H).

Chronic viral co-infections have been associated with an increased likelihood of developing Long COVID and variations in symptom presentation^21^. Additionally, immune system dysregulation attributed to the presence of SARS-CoV-2 reservoirs could also serve as a trigger for the reactivation of latent pathogens^12^. Therefore, we compared the baseline infection history of participants in both arms using Serum Epitope Repertoire Analysis (SERA). The seropositivity (IgG) against a range of pathogens, including six bacterial, eight parasitic, 15 viral, and one fungal species, was evaluated^13,16,22^. No significant differences in seropositivity levels were identified between the two groups (extended data fig. 2).

### No changes in circulating immune cell populations

To determine the impact of treatment on circulating immune cell populations, peripheral blood mononuclear cells (PBMC) were analyzed using flow cytometry (Extended data fig 3A-D). A prior cross-sectional study from our lab has shown significant alterations in specific circulating immune cell populations in individuals with Long COVID compared to convalescent controls^16^. However, no changes in cell populations were observed following NMV/r or PBO/r treatment (extended data fig. 4A-B). Specifically, focusing on the cell types we previously identified as different in Long COVID patients^16^, no differences were observed in non-conventional monocytes (CD14^low^CD16^high^) or conventional type 1 dendritic cells (cDC1; CD304^-^/HLA-DR^+^ /CD141^+^) in either arm at day 28. Similarly, no differences in proportions of activated B lymphocytes (CD86^high^HLA-DR^high^) or double-negative subsets (IgD^-^CD27^−^CD24^−^CD38^−^) were observed upon treatment in either arm. Additionally, no change in proportions of circulating effector memory T lymphocyte populations (CD45RA^−^CD127^+^CCR7^-^) or central memory T cells (CD45RA–CD127+CCR7^+^) were observed. No differences in median percentages of exhausted (PD-1^+^TIM3^+^) CD4^+^ or CD8^+^ subpopulations were observed. Upon stimulation with phorbol myristate acetate and ionomycin, higher, but non-significant, median levels CXCR3 expressing CD4+ cells were observed at day 28 in the NMV/r arm (Paired Wilcoxon Rank Sum test p_adj NMV/r_= 0.067) without corresponding differences of the same in the PBO/r arm (Paired Wilcoxon Rank Sum test p_adj PBO/r_ = 0.9277) (extended data fig. 4C; extended data table 3).

### Treatment alters levels of circulating cytokines in both arms

Numerous studies have identified elevations in markers of systemic inflammation in Long COVID^15,23,24,25^ and a potential association with viral persistence^18^. Additionally, ritonavir has been shown to disrupt endoplasmic reticulum-Golgi protein trafficking, leading to organelle stress responses in the liver^26^. Hence to evaluate potential changes, the concentration of 105 circulating cytokines and hormones were measured using multiplexed Luminex assays. No significant concentration differences were observed both at baseline (Fig 4A; extended data table 1) and at day 28 (Fig 4B; extended data table 1) between the treatment arms. However, more than 30 cytokines were significantly increased or decreased in both the PBO/r (Fig 4C; extended data table 1) and NMV/r (Fig 4C; extended data table 1) arms, with substantial overlap in the specific cytokines affected by day 28 compared to baseline. Among these, a decrease in an inflammatory chemokine RANTES (CCL5), with primary binding specificity to the C-C chemokine receptor type 5 (CCR5)^27^, were observed upon treatment in both study arms and this decrease was further validated by ELISA in both the PBO/r (Wilcoxon rank-sum test; p = 0.0066) and the NMV/r arms (p = 0.0490) (Fig. 4E, F).

**Fig 4:**
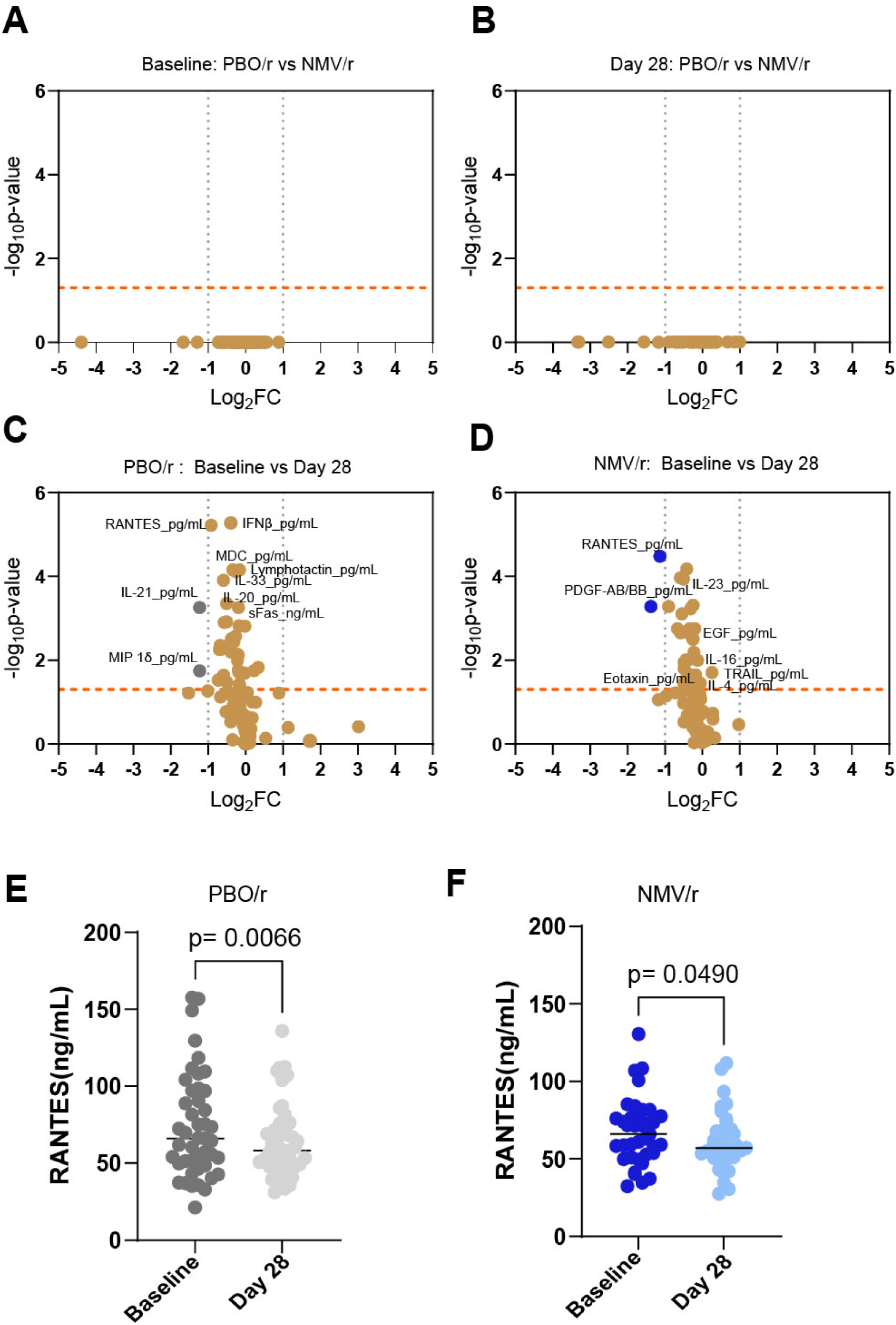
Changes in levels of circulating immune modulators in both the study arms A-B. Volcano plots of differences in baseline and day 28 circulating immune modulators between the two arms of the study. **C-D.** Volcano plots of differences in circulating immune modulators between baseline and day 28 within each arm of the study. The orange dotted line indicates -log_10_p-value of 0.05 and the gray dotted lines mark a negative and positive log_2_fold change of 1. E-F. Shows orthogonal validation of the observed trends in RANTES levels using ELISA. Mann Whitney U Tests were run to evaluate differences between treatment arms. To evaluate differences between timepoints within each treatment arm, Wilcoxon matched-pairs signed-rank test was used. Benjamini Hochberg method of multiple testing correction was implemented.

Both the Stanford trial and our own trial have reported dysgeusia in over 45% of the participants in the NMV/r arm^7,8^, a rate significantly higher than that seen in the 5-day treatment trial among non-immunocompromised participants and in the 15-day trial among immunocompromised participants during the acute phase of infection^28–30^. The likely mediation of bitter taste-sensing type 2 receptor 1 (TAS2R1) in dysgeusia has been reported^31^, which is also known to impact multiple cell types in extraoral organs, regulating the functions including endocrine responses^32^. Hence, we next subdivided the participants in the Paxlovid treatment arm based on reports on dysgeusia to see if there were differences in hormone levels included in this study between the two subsets. Sixteen out of 37 participants in the NMV/r arm had self-reported dysgeusia compared to the 3 out of 45 in the placebo arm, as a treatment-emergent adverse event and significantly higher levels of cortisol were observed among participants who reported dysgeusia (Mann-Whitney U test p_adj_= 0.0414; extended data table 4).

### Decrease in circulating RANTES levels correlates with symptom improvement across treatment arms

Since the 15-day course of NMV/r did not demonstrate efficacy, we sought to identify potential immunological markers of symptom improvement irrespective of treatment arms. Participants who reported improvement in at least four out of five instruments including (PROMIS)-29 v2.1 Physical Health Summary Score (PHSS), PROMIS-29 v2.1 Mental Health Summary Score (MHSS), Modified General Symptom Questionnaire (GSQ)-30, EuroQol EQ-5D-5L Visual Analogue Scale [EQ-VAS], and total symptom burden, were classified as “Improved”. All others were categorized as “non-improved” for this two-group analysis. No significant differences in biological sex (Chi-square test p= 0.5453), age (Mann-Whitney U test p= 0.07), or number of vaccines (Mann-Whitney U test p= 0.86) were observed among the participants in the two groups (Fig. 5A). Differences in circulating immune modulator levels were next evaluated between the two groups. The levels of both IL-5 (Mann-Whitney U test p_adj_= 0.0301) and RANTES (Mann-Whitney U test p_adj_= 0.0319) were observed to be significantly reduced among those who reported alleviation of Long COVID symptoms (Fig. 5B-C). To check for potential confounding by age, sex, and vaccination status, multiple logistic regression models were constructed to assess the contributions of IL-5 and RANTES on improvement status. These models indicated that a decrease in RANTES levels was significantly associated with symptom improvement (p_adj_= 0.0086) whereas IL-5 was no longer significantly associated (p_adj_= 0.291), potentially due to a significant borderline association with age (p_adj_= 0.0686) (Fig. 5D).

**Fig 5.**
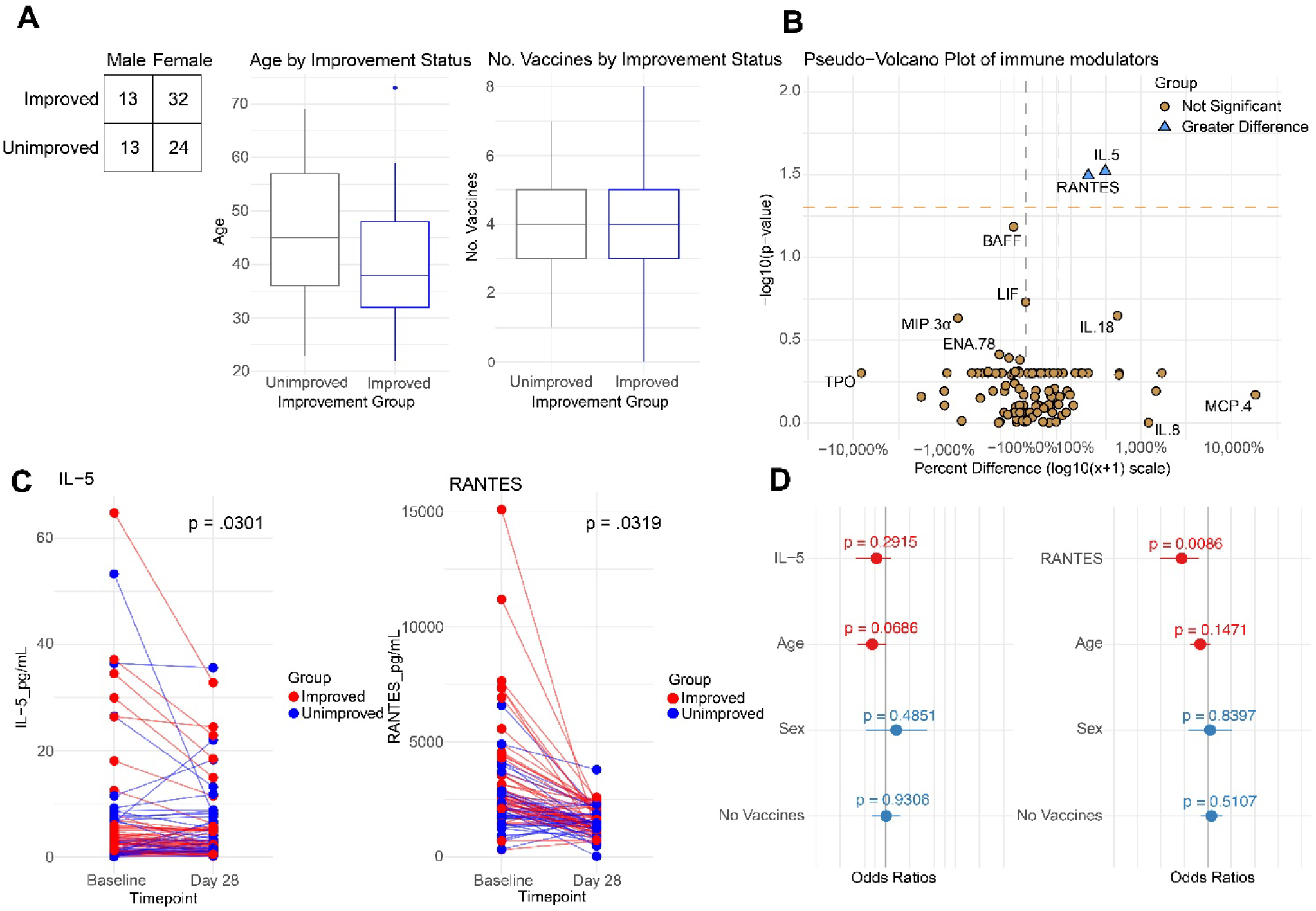
Identification of RANTES as a Potential Improvement Correlate. **A.** Demographic information for improved and unimproved populations. Contingency table with Fisher’s exact test p-value for sex breakdown, and bar plots for the distribution of age and number of vaccines, alongside Mann Whitney U-test p-values. **B.** Pseudo-volcano plot of immune modulators. The x-axis represents percent change from baseline to day 28, with values log transformed for readability. The p-values were calculated using the Mann Whitney U-test with Benjamini Hochberg correction. **C.** Individual-level time-series plots for RANTES and IL-5, alongside corrected p-values. **D.** Logistic regression models predicting improvement status using RANTES or IL-5, alongside age, sex, and number of vaccines.

Beyond univariate comparisons of immune modulators between improvers and non-improvers, we performed unsupervised hierarchical clustering to investigate whether participant groupings based on cytokine profiles recapitulate symptom improvement status. To mitigate the influence of noisy features and preserve meaningful clustering structure, ten immune factors that exhibited the most pronounced differences between improvers and non-improvers were selected for clustering, based on unadjusted p-values from the Wilcoxon test. We then confirmed that the clustering patterns reflected true biological signals rather than artifacts of the feature-selection process by performing a permutation test: we shuffled symptom improvement labels 100 times and re-selected the top 10 modulators in each iteration. The association between the resulting clusters and symptom status in the original data was significantly stronger than expected by chance, indicating that the observed clustering pattern was unlikely to occur by chance (Chi-square test, p = 0.0297). Cluster 2 contained a higher proportion of symptom improvers (66.7%) compared to Cluster 1 (15%), with a significant association between cluster membership and symptom improvement (Chi-square p = 2.0 × 10⁻; permutation test p = 0.0297; extended data fig. 5A). To characterize the overall patterns distinguishing the two clusters, we further performed principal component analysis (PCA), where the first two components (PC1 and PC2) explained 29.6% and 20.4% of the total variance, respectively (extended data fig. 5B). The top contributors to PC1 were LIF, MIP-3β, IL-5, IFNω, and BAFF, while the main contributors to PC2 were RANTES, ENA-78, BAFF, IFNω, and LIF (extended data table 4).

## Discussion

In this exploratory analysis of biological factors before and after treatment with a 15-day course of PBO/r or NMV/r, we made several key observations that inform the clinical outcomes of the PAX LC trial^8^. Participants in each treatment arm were largely comparable at baseline in terms of age, biological sex, viral variant exposure, vaccination history, duration of symptoms, overall health status, and prior exposure to common pathogens. First, we observed no changes in the levels of anti-Spike antibody responses or circulating immune cell subpopulations after treatment. We observed the presence of the Spike protein in circulation in a significant fraction of the participants in both treatment arms; however, no changes in the levels of the Spike protein were observed in either arm. By contrast, both treatment arms exhibited changes in cytokine levels, while modest hematologic changes were observed specifically in the nirmatrelvir arm on day 28. Notably, a decrease in circulating RANTES levels correlated with self-reported symptom improvement across both treatment arms.

None of the Paxlovid trial protocols, including ours, incorporated evidence of viral persistence as a part of participant inclusion criteria. A limitation was that most of these studies used immunoassays that lacked consistency between assays and cohorts. In addition, minimal and intermittent viral shedding made reliable detection of persistent viral antigens from accessible biospecimens challenging^18, 33,34^. We used the SPEAR assay, which is an ultra-sensitive assay based on successive proximity extension and amplification of DNA sequence coupled to a pair of detection antibodies to detect femtomolar levels of target antigens^13^. This enabled baseline detection of SARS-CoV-2 S protein in circulation in a significant subset of participants (∼55% PBO/r, ∼43% NMV/r), enabling the interrogation of the hypothesis of viral persistence. The strong correlation observed between S1 and full-length Spike protein levels, carried out in different batches, supports the specificity and reproducibility of the assay. In contrast, based on the PCR analysis for viral RNA in stool, none of the participants in the Stanford study exhibited positive values at baseline^7^. This difference likely reflects variations in the assay sensitivity, biological target (viral protein vs. viral RNA), sample source (blood vs. stool), and patient population characteristics.

Based on the SPEAR assay, no significant changes in the circulating S1 levels were observed between baseline and day 28, suggesting that NMV/r was ineffective in reducing viral antigens in this Long COVID cohort over the study period. There are a few possible reasons for this outcome. First, we do not understand the molecular nature of the persistent Spike protein *in vivo*. Circulating Spike protein could originate from multiple biological sources. It is possible that the Spike protein is produced in the absence of active viral replication, in which case a 3-chymotrypsin-like cysteine protease (Mpro) inhibitor like NMV/r would not be effective. It is also possible that the residual virus replicates very slowly or sporadically, requiring longer treatment duration to capture an active replication event. However, extended use of Paxlovid may potentially create a resistant virus or toxicity ^35–37^. Another possible reason is the presence of viral reservoirs in inaccessible tissues, where orally administered nirmatrelvir might have limited penetration, enabling viral reservoirs to evade clearance^1^. However, persistent antigenemia, regardless of source, may be biologically relevant in sustaining immune activation in Long COVID.

Nirmatrelvir (PF-07321332) is an inhibitor of Mpro of SARS-CoV-2^6^. After oral administration, it undergoes significant first-pass metabolism in the gastrointestinal tract mediated by cytochrome P450 (CYP) enzymes, and is subsequently further metabolized in the liver predominantly by cytochrome P450 3A4 (CYP3A4).Therefore, ritonavir (Norvir; AbbVie Inc., Chicago, IL, USA), a CYP3A4 inhibitor is co-administered with nirmatrelvir to maintain effective plasma concentrations^38^. Other than CYP3A4, ritonavir is a well-established inhibitor of enzymes and transporters such as P-glycoprotein (P-gp), CYP2D6, and organic anion-transporting polypeptide 1B1 (OATP1B1) among others and can also influence the metabolism of endogenous proteins, peptides^39^. We observed changes in levels of > 30 circulating cytokines in both arms of treatment. While the subtle decreases in these cytokines may have no biological significance, underlying mechanisms warrant further investigation to determine whether they reflect the impact of ritonavir on cellular signaling pathways. The two Paxlovid randomized clinical trial studies ^7,8^ have highlighted dysgeusia and diarrhea as treatment-emergent adverse effects associated with extended NMV/r use. Therefore, the increased levels of cortisol in circulation in participants who reported dysgeusia require further evaluation.

We observed no reduction in anti-Spike IgG levels post-treatment with modest decreases in anti-RBD and anti-N IgG levels by ELISA in both arms. However, anti-N antibody index numbers measured by the COBAS assay showed a slight increase in the placebo arm. PIWAS epitope mapping showed no alterations in anti-S antibody levels against linear peptides upon NMV/r treatment. These data showing no obvious changes in the antibodies against the viral antigens are not surprising, given that the half-life of serum IgG is ∼21 days ^40^, and that we observed no decline in the viral antigen S1 in the participant over time. Further, no significant changes in circulating immune cell subpopulations were observed, other than the borderline increase in the CXCR3^+^ CD4 T cells in the NMV/r arm.

Decreased RANTES levels were associated with symptom improvement, independent of treatment, age, sex, or vaccine status, which merits further exploration in pathophysiology. We do not know whether this association is a simple correlation or causation. The results of two randomized placebo-controlled trials of Maraviroc, a *bona fide* CCR5 antagonist (NCT06974084, NCT06511063) will be informative in understanding whether the CCL5-CCR5 pathway plays a pathological role in Long COVID.

This study has several limitations. The small number of circulating Spike-positive participants may have compromised the robustness of the analyses and reduced the ability to detect subtle treatment-associated changes. Additionally, the absence of a placebo group with no ritonavir and the potential confounding effects of ritonavir could have attenuated the observed changes. Furthermore, given the half-life of serum IgG, the limited follow-up duration may also have contributed to the absence of detectable differences in the treatment arm. Additionally, the 15-day treatment regime may have been too short to fully eliminate potential persistent viral reservoirs. Results from larger Paxlovid trials with longer treatment duration and follow-up period are anticipated to reveal further insights.

In conclusion, although NMV/r was safe and well-tolerated over 15 days, no virological or immunological evidence supports its use as a treatment for Long COVID in this study population. Importantly, this study contributes to guiding future clinical trials exploring alternative durations, dosing, or agents with better tissue penetration. Furthermore, our results underscore the need to expand clinical trials targeting additional pathological drivers of Long COVID, including autoimmunity and chronic inflammation.

## Materials and Methods

### Study design and patient characteristics

The PAX LC study was a decentralized, phase 2, randomized, double-blind, placebo-controlled clinical to investigate the efficacy and safety of a 15-day regimen of orally administered nirmatrelvir/ritonavir compared with placebo/ritonavir in participants with Long COVID. The details of the study has been published elsewhere^8^. Briefly, the key inclusion criteria for participants were: being 18 years or older and documented evidence of previous SARS-CoV-2 infection as confirmed by positive PCR test or medical record of long COVID diagnosis. Key exclusion criteria included acute illnesses such as SARS-CoV-2 infection within the past 2 weeks, active liver disease, renal impairment, or being immunocompromised as defined by the US Centers for Disease Control and Prevention standards. Clinical research coordinators contacted participants via video conference to obtain electronic informed consent using software compliant with the Code of Federal Regulations. The Yale University Institutional Review Board approved this randomized clinical trial (IRB# 2000034086), and the study was conducted under a US FDA investigational new drug application held by Prof. Harlan M Krumholz. This study adhered to the principles of the Declaration of Helsinki and Good Clinical Practice guidelines, was registered with ClinicalTrials.gov (NCT05668091) and followed CONSORT guidelines.

### Biospecimen collection

Whole blood samples were collected in lithium-heparin-coated (BD 367880, BD Biosciences) and sodium-EDTA coated vacutainers (BD 367856, BD Biosciences) from participants’ homes, by ExamOne phlebotomists (Quest Diagnostics) or at the Yale clinic, New Haven, CT. After collecting, biospecimens were either shipped overnight at regulated temperatures or handed over locally to researchers at Yale University in New Haven, CT. Collection tubes were de-identified upon receipt according to protocol and study identifiers were provided. Samples were processed within 48 hours of collection. Biorender^41^ was used to create a graphical schematic of the CONSORT flow chart, study design, cohorts and assays.

At the time of biospecimen collection, the ExamOne phlebotomists were trained to ask the participants to rate their general health status using the following options: excellent, very good, good, fair, poor and do not know and a general health visual analogue scale (GHVAS) having a numerical value between 0 to 100, where higher scores represented better health. These responses were recorded on PAXLC-ExamOne requisition forms and shipped with biospecimens^13^.

### Plasma and PBMC isolation

Previously published protocols were followed for plasma and peripheral blood mononuclear cells (PBMC) isolations^16^. Briefly, whole blood samples from lithium-heparin-coated vacutainers were centrifuged at 650g for 10 minutes at room temperature without braking. Each participant’s plasma was pooled together to a single 15 mL polypropylene conical tube, aliquoted and stored at 80℃. PBMCs were subsequently isolated using SepMate^TM^-50 tubes (StemCell Technologies) following manufacturer’s instructions. Downstream flow analyses were carried out using freshly isolated PBMCs.

### SARS-CoV-2 variant wave determination

SARS-CoV-2 sequence data submitted from the United States were accessed from the GISAID database^11^ on the 14^th^ of January 2025. The dominant viral variants were determined by specifying 6-month class intervals between January 2020 and December 2023. In parallel, the index infection dates reported by participants were classified into concordant time intervals to assign the dominant variants as identified from GISAID. Subsequently, variant waves were assigned based on World Health Organization’s classification system.

### Complete blood counts

An in-house ADVIA hematology Analyzer 2120i (Siemens Healthineers) with 2120i software v 6.11 was used to measure complete blood counts (CBC) from whole blood collected in sodium-EDTA coated vacutainers within 48 hours of collection.

### SARS-CoV-2 Spike Protein Immunoassays

Circulating SARS-CoV-2 S1 levels were measured in plasma samples and validated and by probing for full-length S antigen using Successive Proximity Extension Amplification Reaction (SPEAR) SARS-CoV-2 S protein immunoassays, developed and performed by Spear Bio, Inc. (Woburn, MA) as described previously^13^. Briefly, the assays employ two antibodies with specially designed DNA probes conjugated to two distinct epitopes within the S1 subunit (GenScript, #A02052, #A02058) and both the S1 (GenScript, #A02052) and S2 (R&D Systems, # 979 MAB11362-100) subdomains. The S1 subunit binding antibody was common to both the assays. For the S1 immunoassay, the plasma samples underwent an additional reduction step using proprietary disulfide reduction solution for 15 minutes at 37°C, followed by a quenching step based on plasma processing steps introduced by Swank et al^34^. By contrast, for the full-length Spike assay, denaturation did not alter the assay results and was therefore excluded. The samples were then diluted tenfold in the assay diluent and incubated with S1-specific antibody-oligonucleotide probes for 2 hours at 37°C. This was followed by a 30-minute reaction to convert binding into DNA signal strands which were quantified using quantitative PCR (qPCR) on a QuantStudio 12K Flex platform (Thermo Fisher). Standard curves were generated from a stock solution S1 spike protein spanning from 0.0625 – 100,000 fM. Results from standard curves were used to generate sigmoidal four parameter logistic (4PL) fits in GraphPad Prism (v 10) software. Sample concentration results were calculated from the standard curve of Spike S1 (BioLegend #792904) and full-length Spike (BioLegend #795804), taking dilution factor into account. The LLoD was calculated as the concentration corresponding to 2.5 standard deviation plus mean zero in signal (5 replicates per run). The lower limit of quantification (LLoQ) was defined as the measured concentration at which the curve of measured concentration versus coefficient of variation (CV) intersected at 20% concentration CV. Values below LLoQ were replaced by the assay LLoD value and all values below LLoD were imputed as zero. The values were then transformed using the natural logarithm (ln(x + 1)) for comparisons.

### Immunoglobulin Isotyping

Plasma aliquots were shipped to Eve Technologies on dry ice. To determine the concentrations of the different Ig subtypes and IgG isotypes, the Immunoglobulin Isotyping 6-Plex Custom Luminex Assay (IgG1, IgG2, IgG3, IgG4, lgA, and lgM) (Millipore Sigma, #HGAMMAG-301K) was used.

### SARS-CoV-2 antibody specific immunoassays

The EUA-cleared Elecsys Anti-SARS-CoV-2 immunoassays (Roche Diagnostics, Indianapolis) was used to independently quantitate levels of circulating high affinity anti-N antibodies at the Yale-New Haven hospital clinical research laboratory. The COBAS e801 platform (Roche Diagnostics, Switzerland) was used according to the manufacturer’s instructions as validated earlier ^42^.

Enzyme-linked immunosorbent assays (ELISA) were performed as described earlier^13,16,43^. To summarize, 96-well MaxiSorp plates (Thermo Fisher Scientific, #442404) were coated at a concentration of 2 μg/ml in PBS with recombinant SARS-CoV-2 S protein (50 μl per well; ACROBiosystems, #SPN-C52H9-100 μg) or RBD (ACROBiosystems, #SPD-C52H3-100 μg) or nucleocapsid protein (NUN-C5227-100 μg, ACROBiosystems) and incubated overnight at 4°C. The plates were next blocked with 200 μl of blocking solution (PBS with 0.1% Tween-20 and 3% milk powder) for 1 hour at room temperature (RT) after removing the coating buffer. Plasma samples were diluted 1:1500 (for Anti-Spike and Anti-RBD) and 1:400 (for Anti-N) in buffer (PBS with 0.1% Tween-20 and 1% milk powder). A total of 100 μl of the diluted plasma were added to the wells for 2 hours at room temperature (RT) followed by three washes with PBS-T (PBS with 0.1% Tween-20). A standard curve was generated using serial dilutions of Human anti-Spike [SARS-CoV-2 Human Anti-Spike (AM006415) (Biolegend, #938602)] and anti-nucleocapsid SARS-CoV-2 human anti-nucleocapsid (1A6) (ThermoFisher Scientific, #MA5-35941). A total of 50 μl of horseradish peroxidase conjugated anti-Human IgG antibody (GenScript #A00166; 1:5000) diluted in buffer was added to each well and incubated for an hour. Plates were developed with 100 μl of TMB (3,3’,5,5’-tetramethylbenzidine) Substrate (BD Biosciences, #555214) and absorbance was measured at 450 nm with background correction at 570 nm.

### Linear Peptide Profiling SERA serum screening

A detailed description of the SERA assay has been published^22^. For this study, plasma was incubated with a fully random 12-mer bacterial display peptide library (1 × 10^10^ diversity, 10-fold oversampled) at a 1:25 dilution in a 96-well, deep well plate format. Antibody-bound bacterial clones were selected with 50 µL Protein A/G Sera-Mag SpeedBeads (GE Life Sciences, #17152104010350) (IgG). The selected bacterial pools were resuspended in growth media and incubated at 37 °C shaking overnight at 300 RPM to propagate the bacteria. Plasmid purification, PCR amplification of peptide-encoding DNA and barcoding with well-specific indices was performed as described. Samples were normalized to a final concentration of 4 nM for each pool and run on the Illumina NextSeq500. Every 96-well plate of samples processed for this study contained healthy control run standards to assess and evaluate assay reproducibility and possible batch effects.

### PIWAS analysis

The published^20^ PIWAS method was used to identify antigen and epitope signals against the Uniprot reference SARS-CoV-2 proteome (UP000464024). For each sample, approximately 1–3 million 12-mers were obtained from the SERA assay and these were decomposed into constituent 5- and 6-mers. An enrichment score for each k-mer was calculated by dividing the number of unique 12-mers containing the k-mer divided by the number of expected k-mer reads for the sample, based on amino acid proportions in the sample. A z-score per k-mer was then calculated by comparing the enrichment score with those from a large pre-pandemic cohort (n = 1,500) on a log10 scale. A PIWAS value at each amino acid position along a protein sequence represents an averaged score within a 5 amino acid frame using the tiling z-scores of 5-mers and 6-mers spanning the sequence. 95th quantile bands were calculated based on each population separately.

### Multi-target Luminex assays

Multi-target Luminex assays were used to quantitate levels of circulating immune modulators and hormones. Plasma aliquots stored at −80 °C were shipped to Eve Technologies on dry ice. To avoid batch effects all the samples were run together. The analytes covered by the following panels were assayed: Human Cytokine/chemokine 96-Plex Discovery Assay (HD96), Human Pituitary Panel 1 7-plex (HPTP1), Multispecies Hormone 5-Plex (Millipore Sigma, #MSHMAG-21K). Only analytes with less than 20% missing values were included in the analysis.

Competitive ELISAs were used to quantitate the total plasma testosterone levels. The assays were performed as per manufacturer’s recommendations except for the dissociation time which was increased to 30 mins to ensure complete release of protein bound testosterone (Thermo Fisher, #EIATES).

To validate the total plasma RANTES levels, ELISAs were performed as per manufacturer’s recommendations except for the use of plasma dilution of 1:150 (Thermo Fisher, #EHRNTS). Results from standard curves were used to generate sigmoidal four parameter logistic (4PL) fits in GraphPad Prism (v 10.5.0) software. Sample concentration results were calculated from the standard curve, taking dilution factor into account.

### Flow Cytometry

As described previously, flow cytometry was performed on an Attune NxT Flow Cytometer (Thermo Fisher) using NxT v 5.31.0^13,16^. To summarize, for surface and intracellular staining about 1 to 2 million freshly isolated PBMCs were plated in round-bottom 96 well plates. Dead cells were stained on ice using Live/Dead Fixable Aqua (ThermoFisher), followed by a wash and Fc receptor blocking (Human TruStain FcX™, Biolegend). Surface staining was carried out using a total of 30 fluorophore conjugated antibodies, in three different combinations for surface staining to define myeloid, B and T cell lineages. For quantitation of intracellular cytokines, cells were stimulated by incubating cells in 1× cell stimulation cocktail (eBioscience) without protein transport inhibitor for 1 hour in 10% FBS cRPMI followed by 4-hour incubation in 1x stimulation cocktail with protein transport inhibitor(eBioscience) in 10% FBS cRPMI at 37°C. After stimulation, cells were permeabilized using 1× permeabilization buffer from the FOXP3/Transcription Factor Staining Buffer Set (eBioscience) at 4 °C and stained following Fc receptor blocking. Flow cytometry data were collected with Attune NXT (ThermoFisher) and analyzed using FlowJo v10.8 (BD). The markers used and gating strategies were as described previously ^13,16^ and are detailed at ImmPort portal (Study Registration ID #1776, workspace ID # 7201).

### General Statistical Analyses

Associations between variables between the two treatment arms were analyzed using Chi-square tests for binary variables and the Mann-Whitney U test for continuous variables. Paired comparisons across time points were conducted using the Wilcoxon signed-rank test. Multiple testing correction was applied using the Benjamini-Hochberg method. Spearman’s rank correlation tests were utilized to analyze the relationships between the S protein SPEAR assay values. Adjusted two-sided p-values <0.05 were considered statistically significant, unless otherwise specified. Statistical tests were performed using R (v 4.3.2) and GraphPad PRISM (v 10.5.0).

### Hierarchical clustering and principal component analysis

Hierarchical clustering was performed on scaled data using Euclidean distance and Ward.D2 linkage. The optimal number of clusters was determined by maximizing the average silhouette width across a range of cluster numbers (3 to 8). To refine the effective number of clusters and reduce the influence of unstable small groups, participants in clusters of size ≤ 5 were reassigned to a unified “Grey Cluster”. During permutation, each shuffled dataset was clustered using the same procedure, with the same effective number of clusters retained after isolating the Grey Cluster. The Chi-square statistics measuring the association between cluster membership and symptom improvement status were computed for both the observed and permuted dataset. The permutation p-value was calculated as 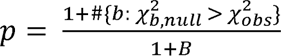, where *B* = 100, and 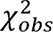 and 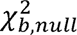 denote the Chi-square statistics from the observed data and b-th permuted dataset, respectively.

We selected the top 10 immune modulators for clustering to capture global patterns, motivated by the observation that 11 modulators had unadjusted p-values < 0.05 in the Wilcoxon test comparing improvers and non-improvers. To avoid potential cherry-picking and assess the robustness of this choice, we evaluated permutation test p-values using different numbers of top-ranked features: top 9 (p = 0.0198), 11 (p = 0.0792), and 15 (p = 0.0891). We did not conduct extensive comparisons with larger feature sets, as including more features tended to reduce cluster separation, likely due to the addition of noise-dominated features with limited discriminatory power.

## Extended data figures

**Extended data fig 1:**
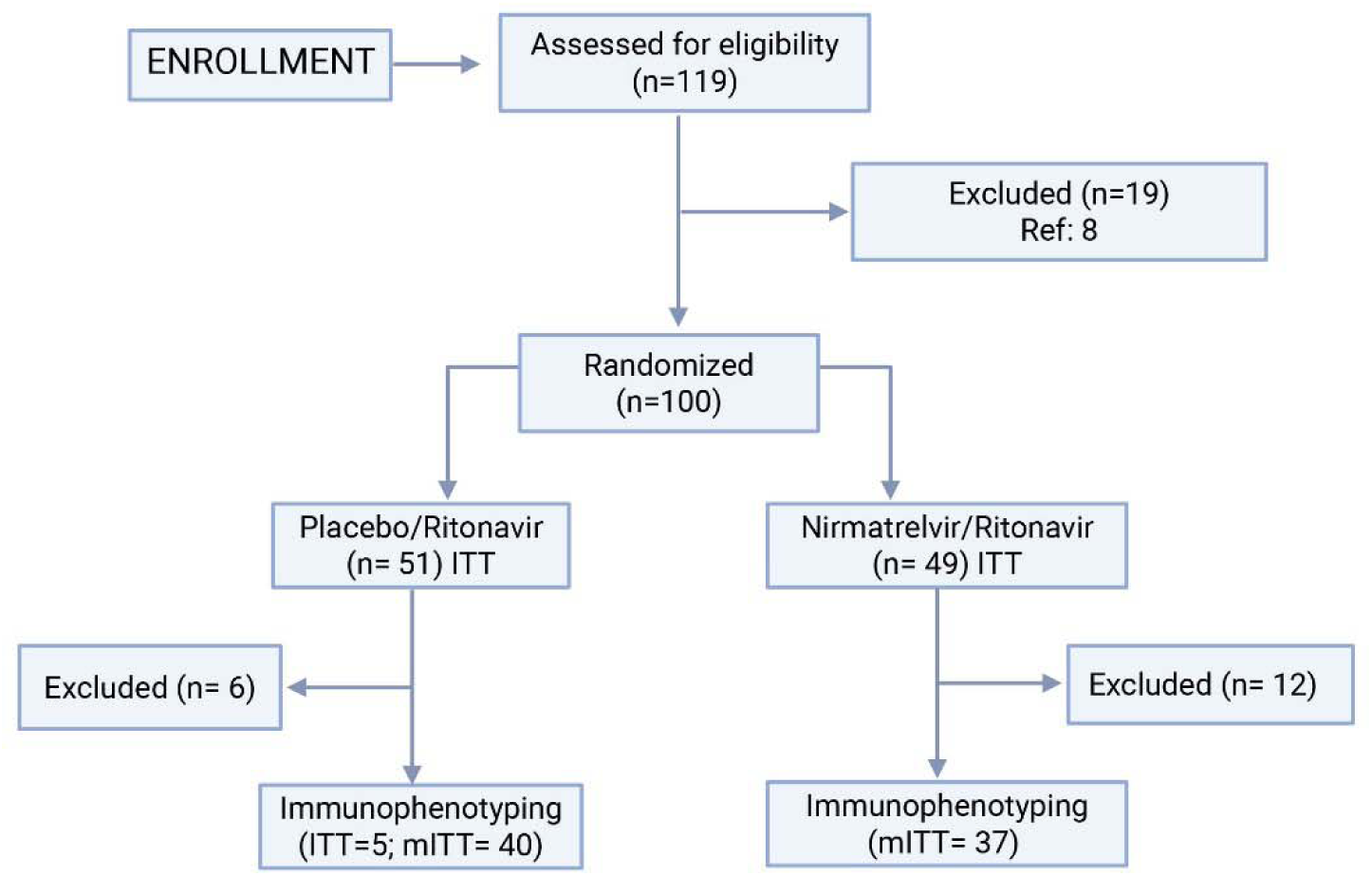
CONSORT Diagram. The terms ITT and mITT indicate the intention to treat population and those with 80% compliance with medications, respectively ^8^. (Among 51 participants in the PBO/r subgroup, 2 withdrew before treatment, 1 was medication incompliant, 1 withdrew consent after the baseline draw, 1 had a failed T1 sample quality assessment, and for 1 the vendor failed to provide phlebotomy services. Among 49 participants in the NMV/r subgroup, 2 did not take medication, 1 had 13% compliance, 3 discontinued on day 5, 1 contracted COVID-19 on day 28, 1 experienced symptom rebound, and 4 had insufficient blood samples despite approved collection attempts.)

**Extended data fig 2:**
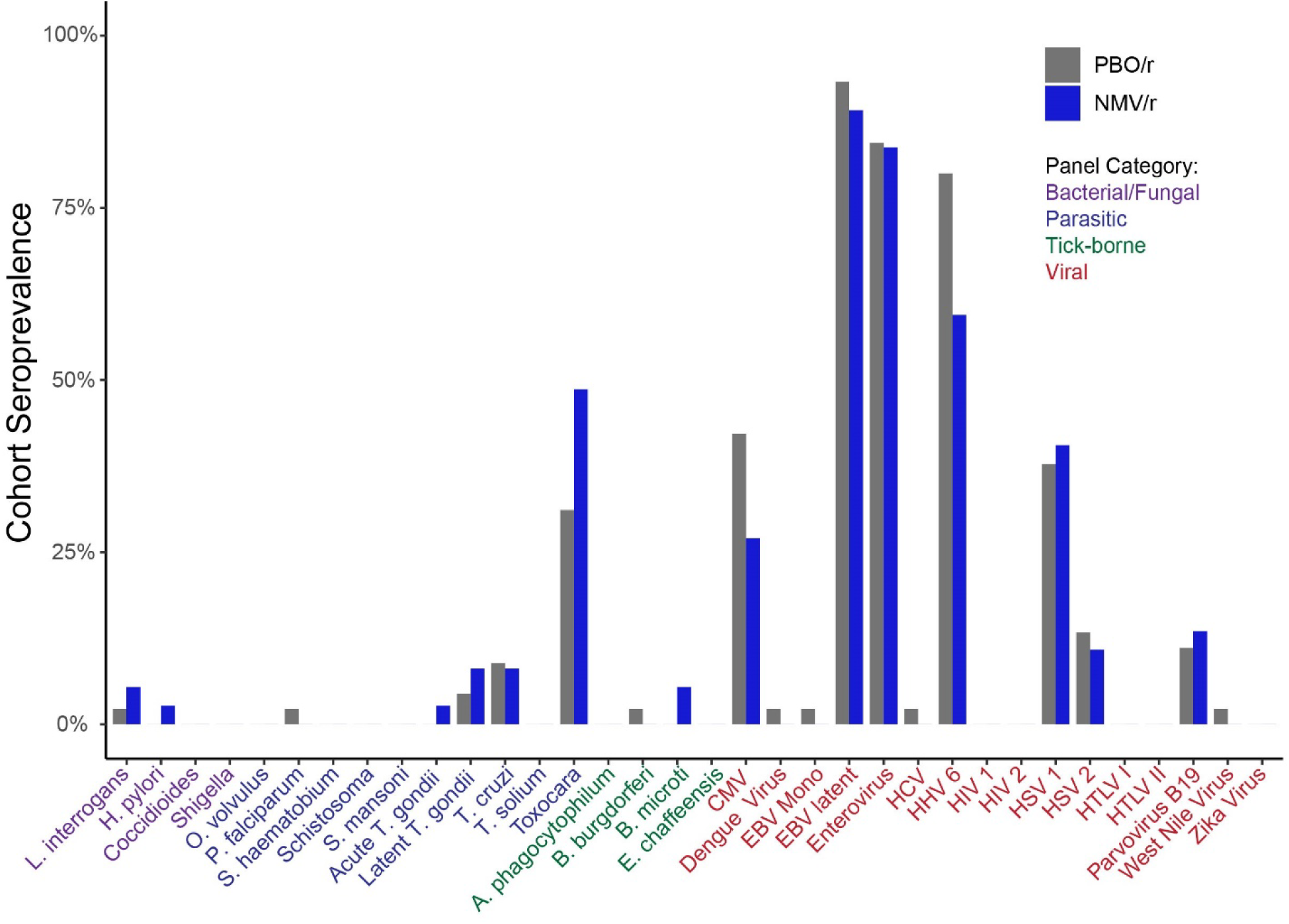
No differences in past exposure to common pathogens between the placebo and treatment groups. The proportion of each group seropositive for each of 34 common pathogen panels as determined by SERA, grouped by pathogen-type. Statistical significance was determined by Fisher’s exact test corrected with FDR (Benjamini Hochberg).

**Extended data fig 3:**
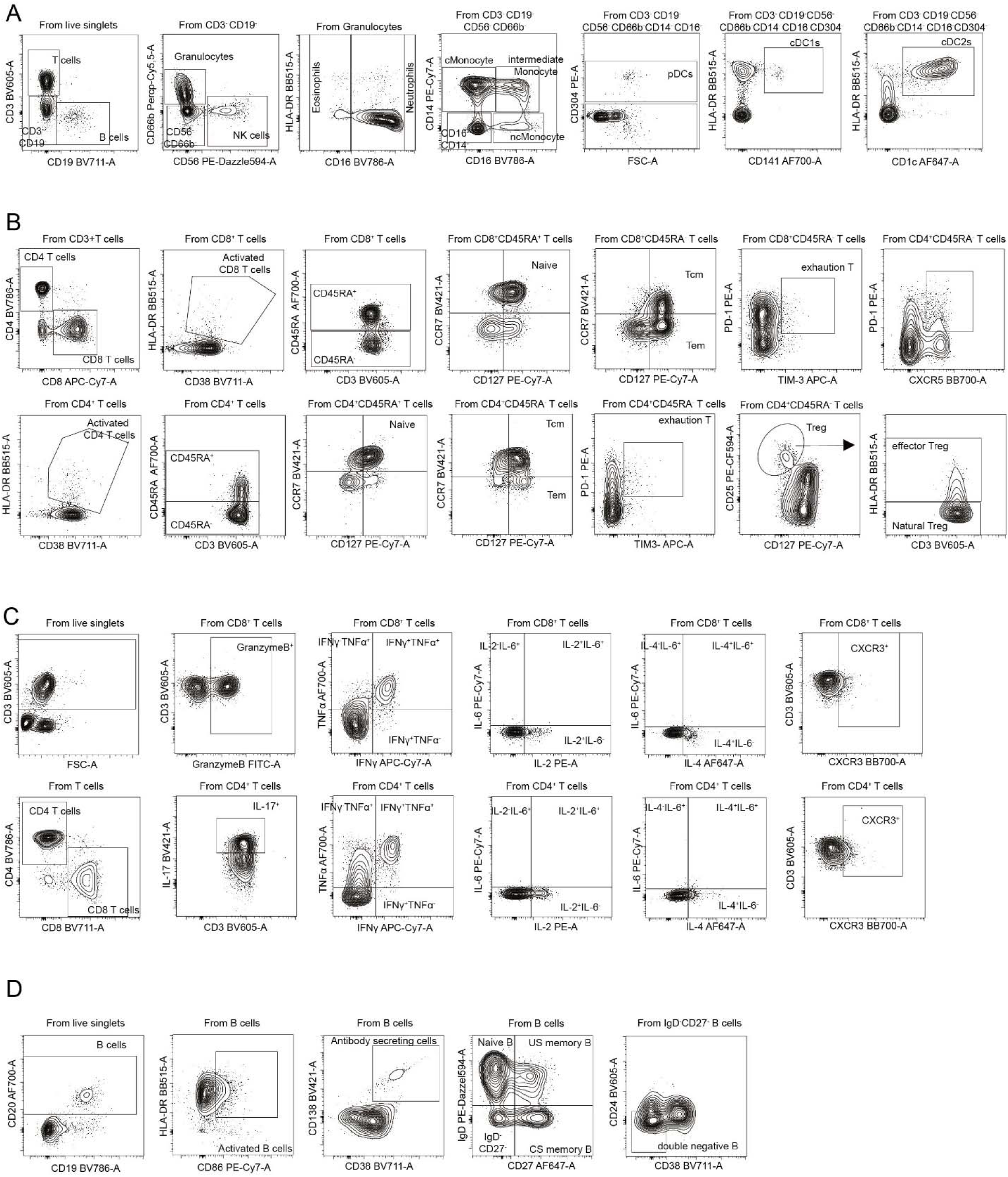
Flow Cytometry gating schematics. **A.** Various gating strategies for granulocyte and myeloid populations. **B.** T lymphocytes **C.** Intracellular cytokine staining and CXCR3 staining **D.** B lymphocytes

**Extended data fig 4:**
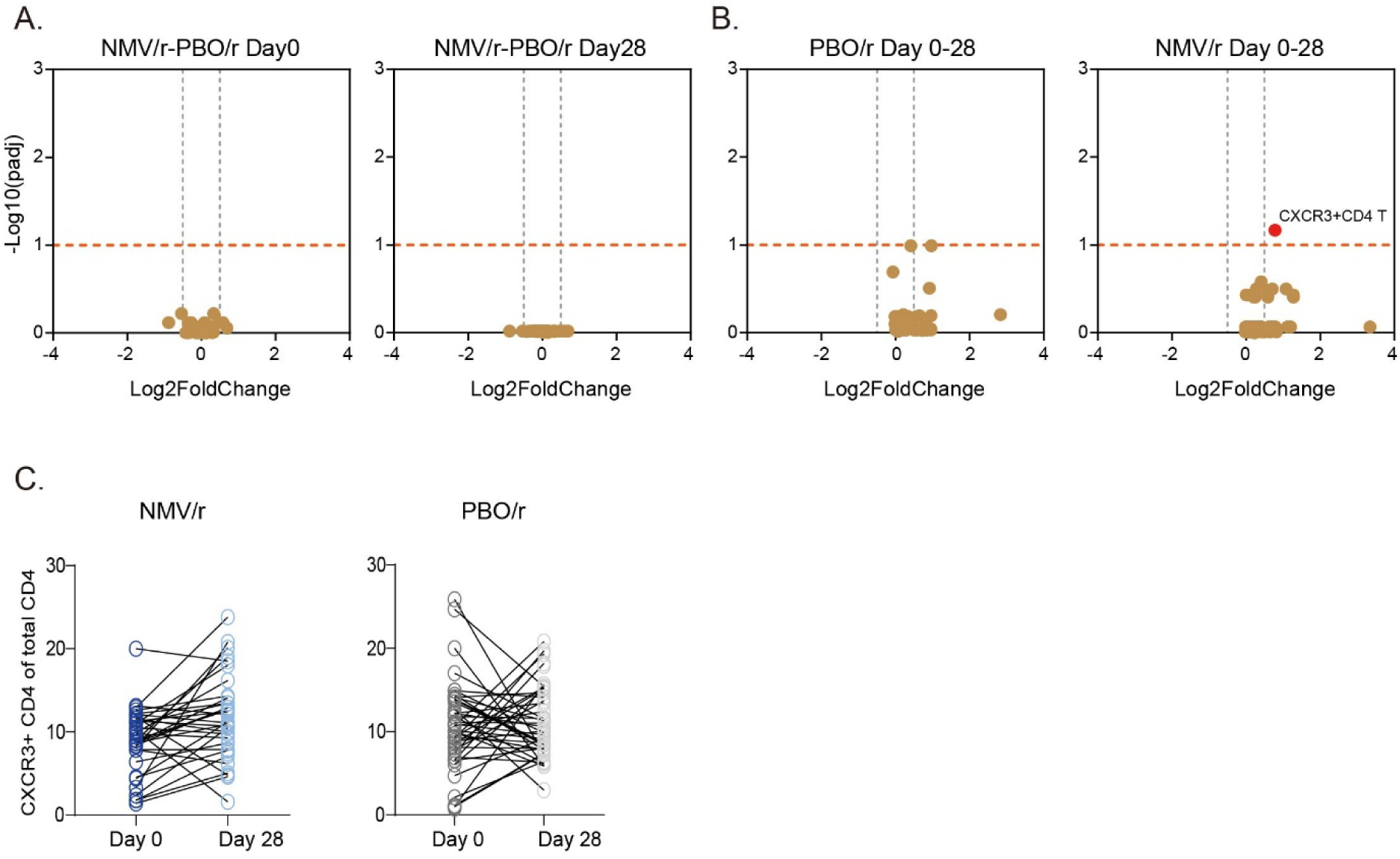
Circulating immune cell population among both study arms. **A.** Volcano plots of difference in baseline and day 28 circulating immune cell populations between the two arms of the study. **B.** Volcano plots of difference in circulating immune cell populations between baseline and day 28 within each arm of the study. Each dot represents an immune cell population including myeloid, B cell, T cell and cytokine producing immune cells. To evaluate differences between timepoints within each treatment arm, Wilcoxon matched-pairs signed-rank test was used. Benjamini Hoochberg method of multiple testing correction was implemented. C. Individual-level time-series plots for CXCR3+ CD4 T cell, alongside corrected p-values.

**Extended data fig 5:**
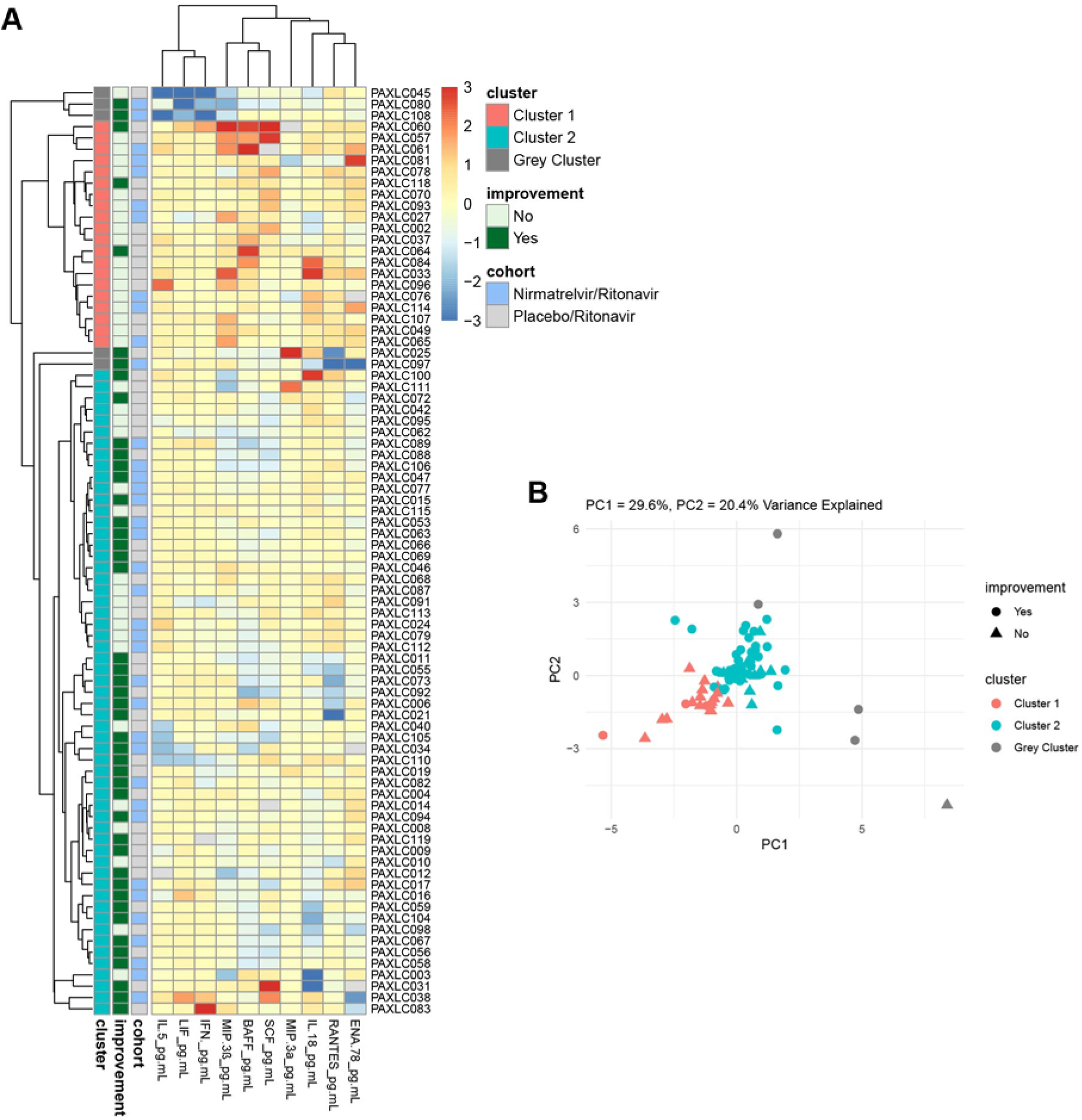
Clustering and PCA of top 10 improvement-associated features: **A**. Eve heatmap of the top 10 features selected by Wilcoxon test to distinguish improvers from non-improvers. Hierarchical clustering was applied to participants, with clusters of ≤ 5 members merged into a unified “Grey Cluster.” A permutation test evaluating the association between improvement status and clustering labels suggests that the observed clustering pattern is not due to chance (p.val = 0.0297). **B**. Principal component analysis of the top 10 features.

## Legends of Extended Data tables

**Extended Data Table 1 |** Demographic, survey and immunological data on all participants included in this study

**Extended Data Table 2 |** Summary of Statistical Analysis results for Complete Blood Count Parameters

**Extended Data Table 3|** Summary of Statistical Analysis results for flow cytometry

**Extended Data Table 4|** Summary of Statistical Analysis on circulating hormones in Paxlovid-treated participants with and without dysgeusia

**Extended Data Table 5 |** Principal Component Loadings and Variable Contributions

## Acknowledgments

We thank Anindita Banerjee, Arthur Bergman, Santos Carvajal-Gonzalez, Robert Fountaine, Jennifer Hammond, Rene Lopez, Amanda Radola, Holly Soares, Brett South, and Erin Stevens from Pfizer for their contributions to this work. This work was supported by funding from Pfizer (grant #76768419). Akiko Iwasaki is an investigator of the Howard Hughes Medical Institute. We thank Melissa Linehan for providing administrative support.

## Author contributions

Experimental conceptualization, methodology and data visualization were performed by B.B., A.T., V.S.M., P.L., G.C.R, V.L.F., K.K. and J.R. for this part of the study. Formal analysis was conducted by B.B., W.B.H., K.W., A.T., V.S.M., PL. Data curation for this part of the study was performed by B.B and W.B.H. Sample processing and biospecimen repository set up were performed by P.B., G.C.R., V.L.F. Clinical investigation and participant management for the PAXLC trial were performed by M.S., B.B., M.A.M, R.K., H.M.K. Statistical analyses plan and formulation for the trial were organized by M.A.M, C.C., H.M.K, S-XL, and J.A.S. M.A.M, B.B., L.C., M.D.J, Y.H., J.Ho, A.C.H, A.I., M.A.J, H.M.K, D.N., E.R., and M.S. were responsible for project administration. B.B., Y.H., J.Ho, A.C.H, A.I., R.K., H.M..K, S-XL, T.B.G, M.S., and F.W.Z. supervised the individuals who were conducting the patient enrolment, eligibility determination, and biospecimen analysis, and supervised all related activities. D.C., Y.H., J.He, S-XL, and M.S. accessed and verified the data for the trial. The original draft was written by B.B. and A.I. Review and editing were performed by B.B., M.S., W.B.H., K.W., A.T., V.S.M., P.L., P.B., G.C.R., V.L.F., K. K., J. R., C.C., R.K., S-XL., J.H., D.C., A.C., F.W., J.H., Y.H., M.A.J., T. B.G., E.R., A.C.H., M.A.M., K.D.C., L.C., M.D.J., D.N., P.A., F.W Z., J. A.S, L.G, H.M.K, A.I. A.I. and H.M.K supervised the study. Funding was acquired by H.M.K. and A.I.

## Data availability

The flow data repository ID will be available for all the raw .fcs files generated for flow cytometry analyses at the ImmPort portal (Study Registration ID #1776, workspace ID # 7201).

## Code availability

R code used for computational analyses will be made available on GitHub.

## References

1 Proal, A. D. et al. SARS-CoV-2 reservoir in post-acute sequelae of COVID-19 (PASC). Nat Immunol 24, 1616–1627 (2023). 10.1038/s41590-023-01601-2

2 Choutka, J., Jansari, V., Hornig, M. & Iwasaki, A. Unexplained post-acute infection syndromes. Nat Med 28, 911–923 (2022). 10.1038/s41591-022-01810-6

3 Davis, H. E., McCorkell, L., Vogel, J. M. & Topol, E. J. Long COVID: major findings, mechanisms and recommendations. Nat Rev Microbiol 21, 133–146 (2023). 10.1038/s41579-022-00846-2

4 Peluso, M. J. & Deeks, S. G. Mechanisms of long COVID and the path toward therapeutics. Cell 187, 5500–5529 (2024). 10.1016/j.cell.2024.07.054

5 Couzin-Frankel, J. Lessons in persistence. Science 384, 150–154 (2024). 10.1126/science.adp7205

6 Owen, D. R. et al. An oral SARS-CoV-2 M(pro) inhibitor clinical candidate for the treatment of COVID-19. Science 374, 1586–1593 (2021). 10.1126/science.abl4784

7 Geng, L. N. et al. Nirmatrelvir-Ritonavir and Symptoms in Adults With Postacute Sequelae of SARS-CoV-2 Infection: The STOP-PASC Randomized Clinical Trial. JAMA Intern Med 184, 1024–1034 (2024). 10.1001/jamainternmed.2024.2007

8 Sawano, M. et al. Nirmatrelvir-ritonavir versus placebo-ritonavir in individuals with long COVID in the USA (PAX LC): a double-blind, randomised, placebo-controlled, phase 2, decentralised trial. Lancet Infect Dis (2025). 10.1016/S1473-3099(25)00073-8

9 Krumholz, H. M. et al. The PAX LC Trial: A Decentralized, Phase 2, Randomized, Double-Blind Study of Nirmatrelvir/Ritonavir Compared with Placebo/Ritonavir for Long COVID. Am J Med 138, 884–892 e884 (2025). 10.1016/j.amjmed.2024.04.030

10 Al-Aly, Z. et al. Long COVID science, research and policy. Nat Med 30, 2148–2164 (2024). 10.1038/s41591-024-03173-6

11 https://gisaid.org/ (accessed 14 January 2025).

12 Proal, A. D. et al. Targeting the SARS-CoV-2 reservoir in long COVID. Lancet Infect Dis 25, e294–e306 (2025). 10.1016/S1473-3099(24)00769-2

13 Bhattacharjee, B., Lu, P., Monteiro, V. S., Tabachnikova, A., Wang, K., Hooper, W. B., Bastos, V., Greene, K., Sawano, M., Guirgis, C., Tzeng, T. J., Warner, F., Baevova, P., Kamath, K., Reifert, J., Hertz, D., Dressen, B., Tabacof, L., Wood, J., Cooke, L., Doerstling, M., Nolasco, S., Ahmed, A., Proal, A., Putrino, D., Guan, L., Krumholz, H. M. & Iwasaki, A. (medRxiv, 2025).

14 Swank, Z. et al. Measurement of circulating viral antigens post-SARS-CoV-2 infection in a multicohort study. Clin Microbiol Infect 30, 1599–1605 (2024). 10.1016/j.cmi.2024.09.001

15 Su, Y. et al. Multiple early factors anticipate post-acute COVID-19 sequelae. Cell 185, 881–895 e820 (2022). 10.1016/j.cell.2022.01.014

16 Klein, J. et al. Distinguishing features of long COVID identified through immune profiling. Nature 623, 139–148 (2023). 10.1038/s41586-023-06651-y

17 Yin, K. et al. Long COVID manifests with T cell dysregulation, inflammation and an uncoordinated adaptive immune response to SARS-CoV-2. Nat Immunol 25, 218–225 (2024). 10.1038/s41590-023-01724-6

18 Rodriguez, L., et al. (medRxiv, 2024).

19 Gao, Y. et al. Identification of soluble biomarkers that associate with distinct manifestations of long COVID. Nat Immunol 26, 692–705 (2025). 10.1038/s41590-025-02135-5

20 Haynes, W. A., Kamath, K., Waitz, R., Daugherty, P. S. & Shon, J. C. Protein-Based Immunome Wide Association Studies (PIWAS) for the Discovery of Significant Disease-Associated Antigens. Front Immunol 12, 625311 (2021). 10.3389/fimmu.2021.625311

21 Peluso, M. J. et al. Chronic viral coinfections differentially affect the likelihood of developing long COVID. J Clin Invest 133 (2023). 10.1172/JCI163669

22 Kamath, K. et al. Antibody epitope repertoire analysis enables rapid antigen discovery and multiplex serology. Sci Rep 10, 5294 (2020). 10.1038/s41598-020-62256-9

23 Cervia-Hasler, C. et al. Persistent complement dysregulation with signs of thromboinflammation in active Long Covid. Science 383, eadg7942 (2024). 10.1126/science.adg7942

24 Liew, F. et al. Large-scale phenotyping of patients with long COVID post-hospitalization reveals mechanistic subtypes of disease. Nat Immunol 25, 607–621 (2024). 10.1038/s41590-024-01778-0

25 Woodruff, M. C. et al. Chronic inflammation, neutrophil activity, and autoreactivity splits long COVID. Nat Commun 14, 4201 (2023). 10.1038/s41467-023-40012-7

26 Khalatbari, A. et al. Ritonavir and Lopinavir Suppress RCE1 and CAAX Rab Proteins Sensitizing the Liver to Organelle Stress and Injury. Hepatol Commun 4, 932–944 (2020). 10.1002/hep4.1515

27 Nelson, P. J. & Krensky, A. M. Chemokines, lymphocytes and viruses: what goes around, comes around. Curr Opin Immunol 10, 265–270 (1998). 10.1016/s0952-7915(98)80164-7

28 Hammond, J. et al. Oral Nirmatrelvir for High-Risk, Nonhospitalized Adults with Covid-19. N Engl J Med 386, 1397-1408 (2022). 10.1056/NEJMoa2118542

29 Cvancara, D. J. et al. Postmarketing Reporting of Paxlovid-Related Dysgeusia: A Real-World Pharmacovigilance Study. Otolaryngol Head Neck Surg 169, 55–61 (2023). 10.1002/ohn.278

30 Weinstein, E. et al. Extended nirmatrelvir-ritonavir treatment durations for immunocompromised patients with COVID-19 (EPIC-IC): a placebo-controlled, randomised, double-blind, phase 2 trial. Lancet Infect Dis (2025). 10.1016/S1473-3099(25)00221-X

31 Caronia, L., Xi, R., Margolskee, R. F. & Jiang, P. Paxlovid mouth likely is mediated by activation of the TAS2R1 bitter receptor by nirmatrelvir. Biochem Biophys Res Commun 682, 138–140 (2023). 10.1016/j.bbrc.2023.10.001

32 Harmon, C. P., Deng, D. & Breslin, P. A. S. Bitter Taste Receptors (T2Rs) are Sentinels that Coordinate Metabolic and Immunological Defense Responses. Curr Opin Physiol 20, 70–76 (2021). 10.1016/j.cophys.2021.01.006

33 Peluso, M. J. et al. Plasma-based antigen persistence in the post-acute phase of COVID-19. Lancet Infect Dis 24, e345–e347 (2024). 10.1016/S1473-3099(24)00211-1

34 Swank, Z. et al. Persistent Circulating Severe Acute Respiratory Syndrome Coronavirus 2 Spike Is Associated With Post-acute Coronavirus Disease 2019 Sequelae. Clin Infect Dis 76, e487–e490 (2023). 10.1093/cid/ciac722

35 Chatterjee, S., Bhattacharya, M., Dhama, K., Lee, S. S. & Chakraborty, C. Resistance to nirmatrelvir due to mutations in the Mpro in the subvariants of SARS-CoV-2 Omicron: Another concern? Mol Ther Nucleic Acids 32, 263–266 (2023). 10.1016/j.omtn.2023.03.013

36 Iketani, S. et al. Multiple pathways for SARS-CoV-2 resistance to nirmatrelvir. Nature 613, 558–564 (2023). 10.1038/s41586-022-05514-2

37 Nooruzzaman, M. et al. Emergence of transmissible SARS-CoV-2 variants with decreased sensitivity to antivirals in immunocompromised patients with persistent infections. Nat Commun 15, 7999 (2024). 10.1038/s41467-024-51924-3

38 Toussi, S. S. et al. Pharmacokinetics of Oral Nirmatrelvir/Ritonavir, a Protease Inhibitor for Treatment of COVID-19, in Subjects With Renal Impairment. Clin Pharmacol Ther 112, 892–900 (2022). 10.1002/cpt.2688

39 Hendrick, V. et al. Pharmacovigilance of Drug-Drug Interactions with Nirmatrelvir/Ritonavir. Infect Dis Ther 13, 2545–2561 (2024). 10.1007/s40121-024-01050-w

40 Booth, B. J. et al. Extending human IgG half-life using structure-guided design. MAbs 10, 1098–1110 (2018). 10.1080/19420862.2018.1490119

41 Scientific Image and Illustration Software-Biorender. https://www.biorender.com/.

42 El-Khoury, J. M., Schulz, W. L. & Durant, T. J. S. Longitudinal Assessment of SARS-CoV-2 Antinucleocapsid and Antispike-1-RBD Antibody Testing Following PCR-Detected SARS-CoV-2 Infection. J Appl Lab Med 6, 1005–1011 (2021). 10.1093/jalm/jfab030

43 Filardi, B. A. et al. Age-dependent impairment in antibody responses elicited by a homologous CoronaVac booster dose. Sci Transl Med 15, eade6023 (2023). 10.1126/scitranslmed.ade6023

